# Long-term trends in stroke and the impact of population aging in China: Results from the 2021 Global Burden of Disease Study

**DOI:** 10.64898/2026.01.29.26345177

**Authors:** Huifeng Liang, Wei Wei, Mihriya Mutallip, XiaoCheng Bao, ShuQun Yang, Chun Zhang, Xin Chen

**Author notes:** **Correspondence:** Corresponding Author: Xin Chen, PhD, Email address, Address: Dalian Medical University, No. 9, West Section of Lvshun South Road, Dalian, Liaoning, China.

## Abstract

**Background:** Stroke is a leading cause of public health burden in China, particularly among the elderly. This study aims to examine long-term trends in stroke incidence and the impact of population aging.

**Methods:** Using the Global Burden of Disease (GBD) Study 2021, we analyzed the incidence, mortality, and disability-adjusted life years (DALYs) for ischemic stroke (IS), cerebral hemorrhage (ICH), and subarachnoid hemorrhage (SAH) from 1990 to 2021. We applied the estimated annual percentage change (EAPC) and decomposition analysis to assess trends and the influence of population aging.

**Findings:** From 1990 to 2021, the age-standardized incidence rate (ASIR) of IS rose from 110.05 to 135.79, with an EAPC of 0.94. The EAPCs for ICH and SAH were –2.24 and –3.70, respectively. Population aging significantly contributed to the stroke burden, with 800,000 IS-related deaths from 1980 to 2021. In 2021, the proportion of IS deaths due to aging was 279.4% for men and 204.8% for women.

**Conclusions:** Stroke incidence and mortality continue to rise, especially among the elderly. Aging exacerbates the stroke burden, highlighting the need for targeted policies to improve the quality of life for the aging population.

## Introduction

Stroke is a leading cause of morbidity, death and disability, with a high disease burden, especially in some low-income and developing countries^[1, 2]^. China has been one of the most representative developing countries, and the results of the Global Burden of Disease (GBD) study show that the number of deaths caused by stroke in China in 2021 was about 2.59 million, accounting for about one-third of the world’s stroke deaths each year, including ischemic stroke (IS), intracerebral hemorrhage (ICH) and subarachnoid hemorrhage (SAH). Stroke is now a growing threat to us and a major public health problem^[3]^.

In recent decades, the epidemiological characteristics of stroke in China have changed significantly^[4]^. China is one of the fastest-aging countries in the world and is facing many issues related to population aging. The world has the highest number of people aged 65 years and above, and by 2050, this demographic is expected to account for 25% of the total population^[5]^. With the increasing age of the Chinese population, the incidence of stroke is likely to rise further. The elderly are at high risk of stroke, and the older the age, the higher the probability of stroke. The decline in physical function and the increase in chronic diseases among older people have created problems for national health and social security systems^[5]^. The burden of stroke disease in China has been analyzed in the published literature, but there is a lack of systematic analysis of stroke subtypes with respect to China’s long-term aging population, which may restrict the ability of policy makers to reform the health care system and lead them to fail to adequately address the healthcare needs of the elderly.

This study is based on data from the latest GBD study (2021). We analyzed incidence and disability indicators from 1990 to 2021 and related death indicators from 1980 to 2021, and then conducted age standardization and gender analysis, respectively. The estimated annual percentage change (EAPC) was calculated to describe long-term trends in stroke in China. We also used a decomposition method to estimate the impact of population aging on stroke^[6, 7]^, to assess the relationship between changes in stroke and population aging in China in recent decades. This analysis provides a scientific basis for more effectively addressing the disease management challenges posed by an aging population.

## Methods

### Data sources

The GBD 2021 provided an assessment of 369 diseases and injuries across 204 countries and territories between 1990 and 2021^[8]^. The details of the methods and data used in the GBD 2021 are described on the Global Health Data Exchange GHDx tool (http://ghdx.healthdata.org/gbd-results-tool). We collected stroke incidence, mortality, and disability-adjusted life years (DALYs) data for China from the database. The primary sources for the original data used in GBD 2021 included the Cause of Death Reporting System of the Chinese Center for Disease Control and Prevention, Disease Surveillance Points, and the Maternal and Child Surveillance System, which are recognized as nationally representative^[8]^.

The socio-demographic index (SDI) is a comprehensive index that measures the population structure and composition of a region. The value of the SDI ranges from 0 to 1. Areas with SDI closer to 1 are shown to be more consistent with healthy development, higher income, longer schooling, and lower fertility rate. Regions with SDI near 0 have the lowest development level, in theory. The 204 countries and territories were categorized into five groups based on their SDI quintile: Low, Low-middle, Middle, High-middle, and High SDI^[9]^.

The search strategies of the selected data were as follows: location name was “China”; the causes were “Stroke,” “Ischemic stroke”, “Intracerebral hemorrhage”, and “Subarachnoid hemorrhage”; measures were “Incidence”, “Deaths” and “DALYs”; and sexes were “Both”, “Male”, and “Female”, We divided the data into 20 age groups of 5 years. The 95% uncertainty intervals (95% UIs) were utilized to compute these indicators.

### Stroke definition

A stroke is an abrupt cerebrovascular incident typically caused by the rupture or blockage of a blood vessel, resulting in the interruption of blood supply to a portion of the brain, resulting in nerve cell damage and dysfunction. According to the World Health Organization, stroke may be divided into three subtypes, IS, ICH, and SAH^[10]^. IS is caused by blockage of an artery, leading to a lack of blood supply to the brain. ICH is characterized by a ruptured blood vessel inside the brain that leaks blood into the brain tissue. SAH involves leakage of blood into the subarachnoid space of the brain, usually caused by a ruptured brain aneurysm or other vascular abnormality^[11]^. There was no detailed classification in GBD 2021^[4]^.

### Statistical analysis

Stroke incidence, deaths, and DALYs in China were analyzed by subtype, sex, and age. Overall trends in age-standardized incidence rate (ASIR) and age-standardized disability-adjusted life-year rate (ASDR) were measured in 20 age groups from 1990 to 2021, and age-standardized mortality rate (ASMR) from 1980 to 2021. The EAPC was used to assess long-term trends in ASIR, ASMR, and ASDR; the EAPC and 95% confidence interval (95%CI) were calculated using the log-linear regression equation, as described previously ^[12]^. The 95%CI was also derived from the fitted model. When EAPC and its 95%CI were greater than 0, the age-standardized rate increased. When it was less than 0, the age-standardized rate decreased. If the 95% CI was 0, the age-standardized rate remained stable.

To find the trend change points of the data over time, Joinpoint regression analysis was used. This analysis method is a statistical technique used to explore potential points of change in data, and its basic principle is to simulate changes in the data by fitting multiple linear segments. By fitting different regression lines on these segment points, it is possible to find the points that best match the change in the data and determine the trend of change in the data.

Decomposition analysis was used in our study to attribute differences in incidence, death, and DALYs between two time points at a site to three independent factors, (a) population size, (b) population aging, and (c) age-specific rates. Age-specific rates demonstrate the impact of factors beyond population size and aging of the population. The M_p_, M_s_, and M_m_ represent the main effects of population size and population aging, and the change of age-specific rates. The I_ps_, I_pm_, and I_sm_ represent the pairwise interactions among the three factors, and I_psm_ represents the interaction among the three factors. The specific formula is as follows^[6, 7]^:

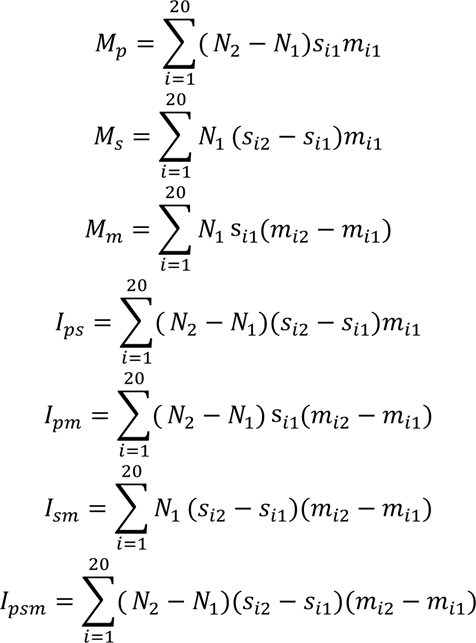

The difference between the two time points can be attributed to changes in population growth (P), population aging (S), and age-specific rates (M), based on the following formula to calculate the contribution of each factor:

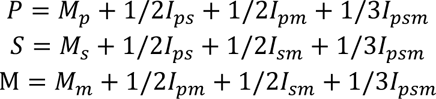

The decomposition analysis of morbidity and disability in the study was based on 1990 as a reference, and the attribution ratio was calculated as the number of deaths and DALYs caused by population aging divided by the total number in 1990. The proportion of deaths attributable to mortality was based on 1980 data. If the number of attributions in the current year exceeded the total number in 1990, it could have exceeded 100%. A positive indicator signifies an elevation in stroke-related metrics, whereas a negative indicator denotes a decline in stroke-related metrics. This analysis, which accounted for differences across time, is critical for supporting decision-making to improve public health at all levels.

All statistical analyses were performed using R4.3.1 and Joinpoint Regression Program (version 5.1.0).

## Results

### Long-term trends of stroke in China

During the past 30 years, the number of stroke cases in China increased, while the ASIR showed a downward trend. For IS, there were 2772.05 thousand cases (95%UI: 2295.71, 3319.15), and the ASIR per 100,000 population increased from 100.05 in 1990 to 135.79 in 2021 (EAPC, 0.94 [95%CI, 0.88 to 1.00]). For ICH, there were 1173.29 thousand cases (95%UI: 1003.99, 1330.46), and the ASIR per 100,000 population decreased from 108.93 in 1990 to 61.15 in 2021 (EAPC, –2.24 [95%CI, –2.5 to –1.98]). For SAH, there were 145.14 thousand cases (95%UI: 125.43, 169.02), and the ASIR per 100,000 population decreased from 17.96 in 1990 to 7.81 in 2021 (EAPC, –3.70 [95%CI, –4.08 to –3.31]) (Table 1). In 2021, IS resulted in 1176.95 thousand deaths (95%UI: 986.88, 1372.71) and an ASMR of 64.47 per 100,000 people (95%UI: 54.03, 74.82) in the total population of China (EAPC, –0.34 [95%CI, –0.49 to –0.18]) (Table 2). In 2021, IS resulted in 23,430.41 thousand DALYs (95%UI: 19,918.85, 26,933.91) and an ASDR of 1180.98 per 100,000 people (95%UI: 1009.70, 1356.67) (EAPC, –0.5 [95%CI, –0.69 to –0.32]) (Table 3). Male and female individuals exhibited similar incidence, mortality, and DALY rates. The EAPC of both ICH and SAH showed a negative trend, but the decline of SAH was more obvious than that of ICH (Tables 1–3).

**Table 1.**
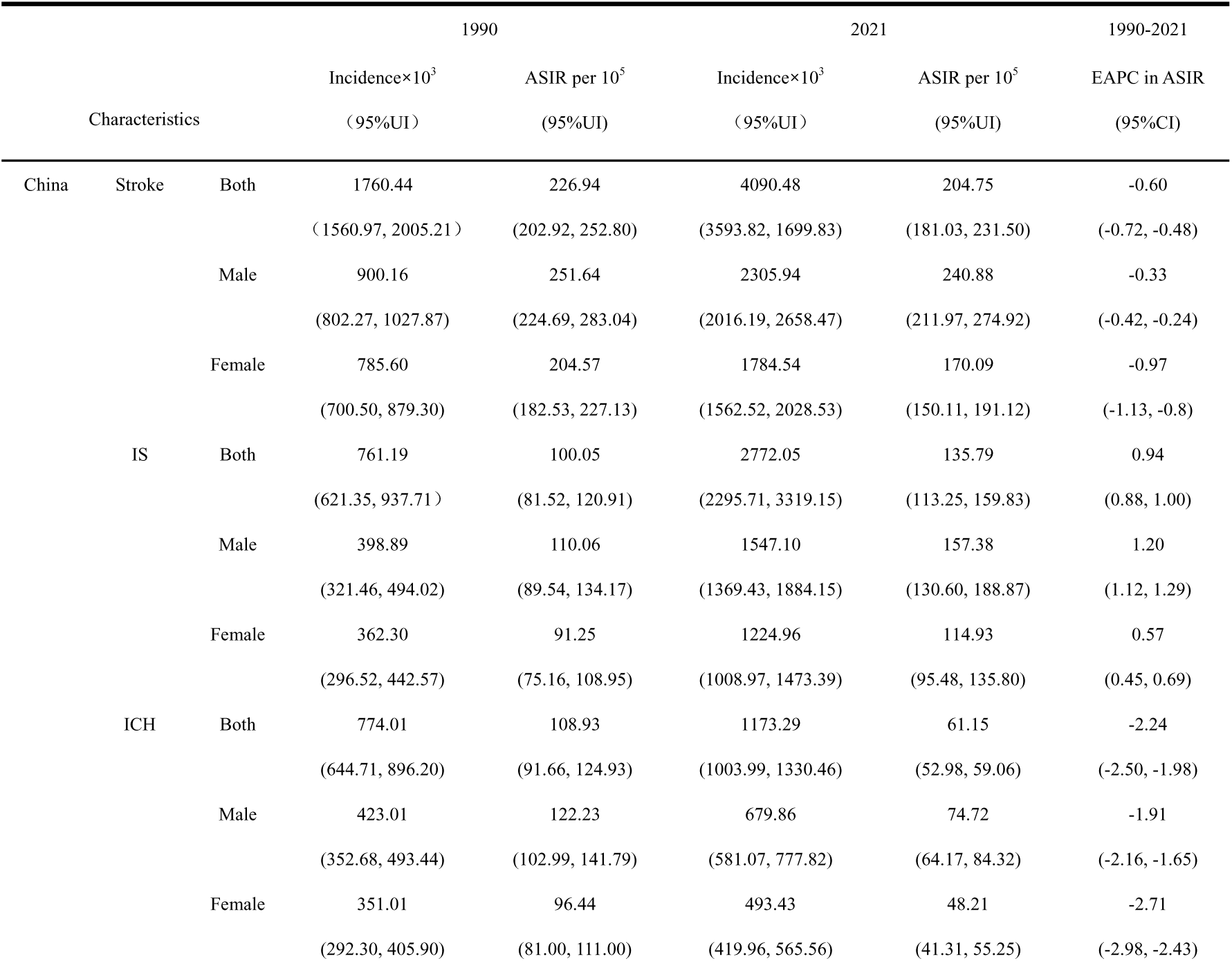
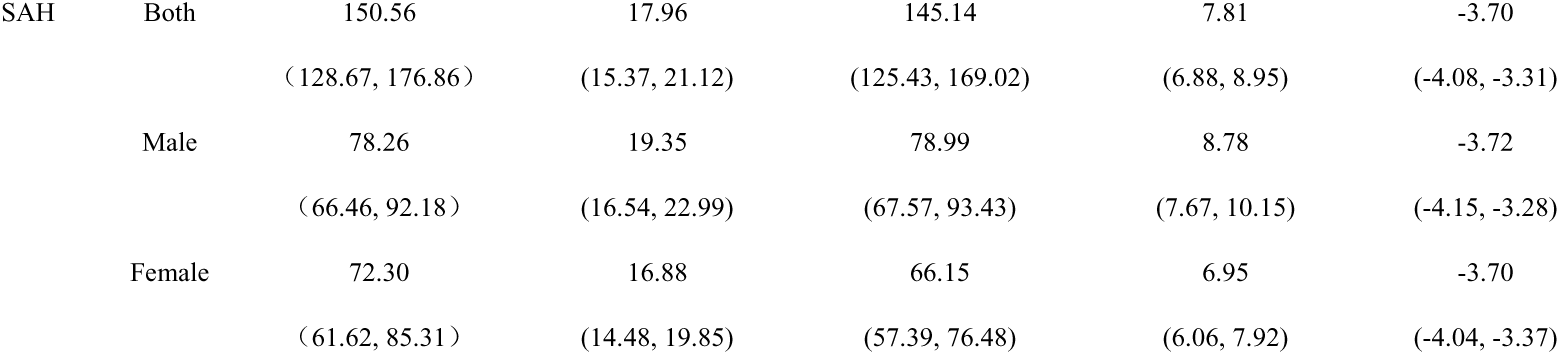
Subtype analysis of stroke cases, incidence and EAPC from 1990 to 2021 in China.

|  |  |  | 1990 |  | 2021 |  | 1990-2021 |
| --- | --- | --- | --- | --- | --- | --- | --- |
| Characteristics | | | Incidence$\times 10^3$<br>(95%UI) | ASIR per $10^5$<br>(95%UI) | Incidence$\times 10^3$<br>(95%UI) | ASIR per $10^5$<br>(95%UI) | EAPC in ASIR<br>(95%CI) |
| China | Stroke | Both | 1760.44<br>(1560.97, 2005.21) | 226.94<br>(202.92, 252.80) | 4090.48<br>(3593.82, 1699.83) | 204.75<br>(181.03, 231.50) | -0.60<br>(-0.72, -0.48) |
|  |  | Male | 900.16<br>(802.27, 1027.87) | 251.64<br>(224.69, 283.04) | 2305.94<br>(2016.19, 2658.47) | 240.88<br>(211.97, 274.92) | -0.33<br>(-0.42, -0.24) |
|  |  | Female | 785.60<br>(700.50, 879.30) | 204.57<br>(182.53, 227.13) | 1784.54<br>(1562.52, 2028.53) | 170.09<br>(150.11, 191.12) | -0.97<br>(-1.13, -0.8) |
|  | IS | Both | 761.19<br>(621.35, 937.71) | 100.05<br>(81.52, 120.91) | 2772.05<br>(2295.71, 3319.15) | 135.79<br>(113.25, 159.83) | 0.94<br>(0.88, 1.00) |
|  |  | Male | 398.89<br>(321.46, 494.02) | 110.06<br>(89.54, 134.17) | 1547.10<br>(1369.43, 1884.15) | 157.38<br>(130.60, 188.87) | 1.20<br>(1.12, 1.29) |
|  |  | Female | 362.30<br>(296.52, 442.57) | 91.25<br>(75.16, 108.95) | 1224.96<br>(1008.97, 1473.39) | 114.93<br>(95.48, 135.80) | 0.57<br>(0.45, 0.69) |
|  | ICH | Both | 774.01<br>(644.71, 896.20) | 108.93<br>(91.66, 124.93) | 1173.29<br>(1003.99, 1330.46) | 61.15<br>(52.98, 59.06) | -2.24<br>(-2.50, -1.98) |
|  |  | Male | 423.01<br>(352.68, 493.44) | 122.23<br>(102.99, 141.79) | 679.86<br>(581.07, 777.82) | 74.72<br>(64.17, 84.32) | -1.91<br>(-2.16, -1.65) |
|  |  | Female | 351.01<br>(292.30, 405.90) | 96.44<br>(81.00, 111.00) | 493.43<br>(419.96, 565.56) | 48.21<br>(41.31, 55.25) | -2.71<br>(-2.98, -2.43) |

|  |  |  |  |  |  |  |
| --- | --- | --- | --- | --- | --- | --- |
| SAH | Both | 150.56 | 17.96 | 145.14 | 7.81 | -3.70 |
|  |  | (128.67, 176.86) | (15.37, 21.12) | (125.43, 169.02) | (6.88, 8.95) | (-4.08, -3.31) |
|  | Male | 78.26 | 19.35 | 78.99 | 8.78 | -3.72 |
|  |  | (66.46, 92.18) | (16.54, 22.99) | (67.57, 93.43) | (7.67, 10.15) | (-4.15, -3.28) |
|  | Female | 72.30 | 16.88 | 66.15 | 6.95 | -3.70 |
|  |  | (61.62, 85.31) | (14.48, 19.85) | (57.39, 76.48) | (6.06, 7.92) | (-4.04, -3.37) |

**Table 2.** Subtype analysis of stroke deaths, mortality and EAPC from 1980 to 2021 in China.

|  |  |  | 1980 |  | 2021 |  | 1980-2021 |
| --- | --- | --- | --- | --- | --- | --- | --- |
|  |  |  | Death×10<sup>3</sup> | ASMR per 10<sup>5</sup> | Death×10<sup>3</sup> | ASMR per 10<sup>5</sup> | EAPC in ASMR |
| Characteristics |  |  | (95%UI) | (95%UI) | (95%UI) | (95%UI) | (95%CI) |
| China | Stroke | Both | 1337.46 | 286.54 | 2591.65 | 138.03 | -1.67 |
|  |  |  | (1150.84, 1556.42) | (251.38, 327.64) | (2179.40, 3032.70) | (116.69, 160.32) | (-1.82, -1.51) |
|  |  | Male | 644.04 | 318.50 | 1506.01 | 186.83 | -1.11 |
|  |  |  | (513.57, 798.90) | (262.62, 380.93) | (1216.25, 1855.99) | (152.41, 226.91) | (-1.28, -0.93) |
|  |  | Female | 693.42 | 266.91 | 1085.64 | 103.71 | -2.27 |
|  |  |  | (558.14, 838.73) | (219.13, 319.37) | (854.44, 1336.35) | (81.60, 127.59) | (-2.43, -2.11) |
|  | IS | Both | 334.86 | 81.60 | 1176.95 | 64.47 | -0.34 |
|  |  |  | (280.24, 406.30) | (69.01, 98.18) | (986.88, 1372.71) | (54.03, 74.82) | (-0.49, -0.18) |
|  |  | Male | 154.57 | 90.89 | 674.87 | 88.32 | 0.20 |
|  |  |  | (115.17, 209.40) | (68.39, 119.71) | (538.08, 831.21) | (71.73, 107.19) | (0.03, 0.38) |
|  |  | Female | 180.29 | 76.24 | 502.09 | 48.58 | -0.90 |
|  |  |  | (144.09, 222.06) | (62.02, 93.17) | (395.76, 616.66) | (38.15, 59.77) | (-1.06, -0.74) |
|  | ICH | Both | 837.03 | 173.12 | 1322.89 | 68.84 | -2.06 |
|  |  |  | (709.63, 993.21) | (147.01, 204.00) | (1108.05, 1567.71) | (57.61, 81.17) | (-2.28, -1.85) |
|  |  | Male | 411.40 | 193.74 | 782.06 | 92.84 | -1.54 |
|  |  |  | (313.18, 524.29) | (152.21, 239.76) | (614.98, 972.67) | (74.42, 113.29) | (-1.77, -1.31) |
|  |  | Female | 425.62 | 159.89 | 540.83 | 51.11 | -2.67 |
|  |  |  | (334.71, 545.49) | (127.21, 201.52) | (419.64, 675.01) | (39.70, 63.79) | (-2.89, -2.44) |
|  | SAH | Both | 165.58 | 31.82 | 91.80 | 4.72 | -5.67 |
|  |  |  | (81.52, 243.52) | (14.97, 47.49) | (66.67, 116.22) | (3.45, 5.95) | (-6.15, -5.18) |
|  |  | Male | 78.07 | 33.86 | 49.09 | 5.68 | -5.13 |
|  |  |  | (25.79, 126.29) | (10.44, 55.59) | (29.04, 69.32) | (3.31, 7.93) | (-5.55, -4.70) |
|  |  | Female | 87.51 | 30.78 | 42.72 | 4.02 | -6.21 |
|  |  |  | (38.96, 129.03) | (13.35, 45.66) | (29.21, 57.85) | (2.76, 5.43) | (-6.75, -5.67) |

**Table 3.**
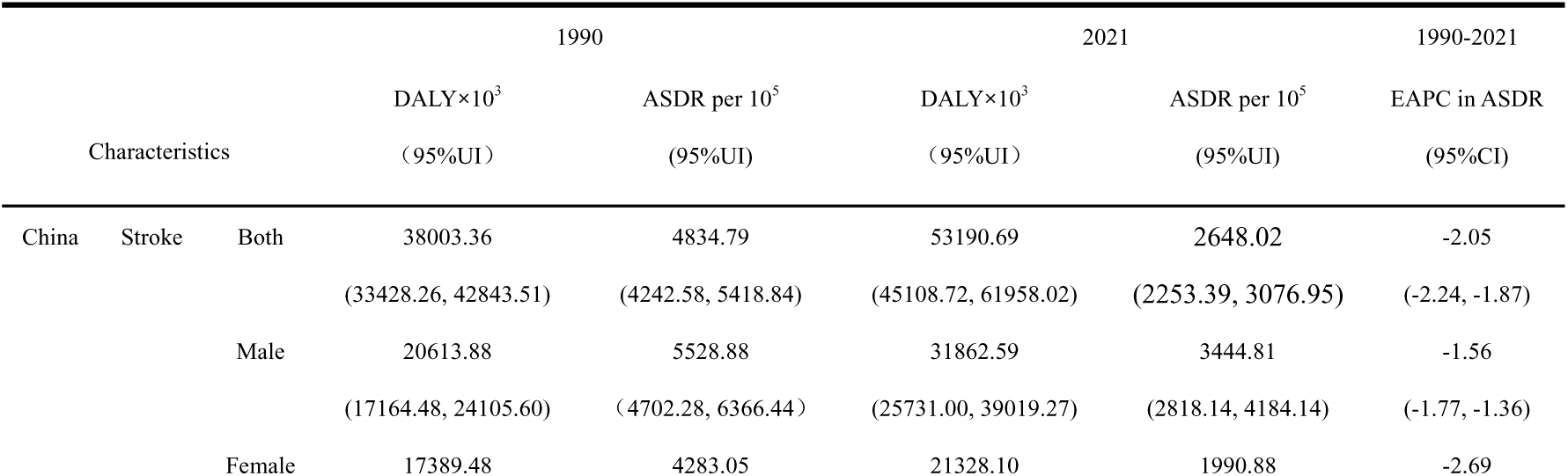
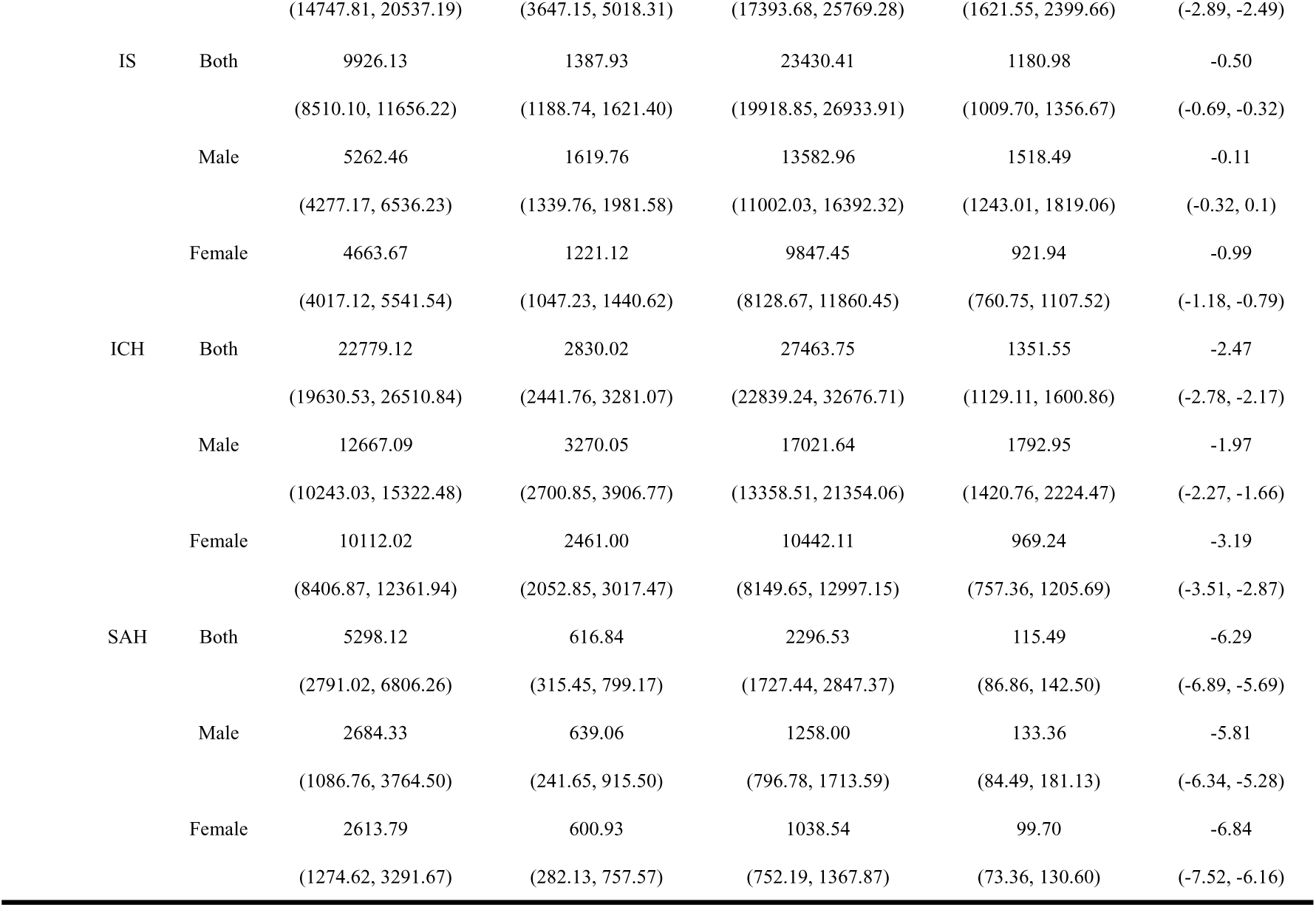
Subtypes analysis stroke of DALYs, DALY rates and EAPC from 1990 to 2021 in China.

From 1990 to 2021, the incidence of ischemic stroke (IS) among Chinese men over 10 years old has been increasing, with the most obvious trend between 75 and 79 years of age (EAPC, 1.88 [95%CI, 1.73 to 2.02]), while the incidence was rising in women over 35 years old. It is worth noting that the DALYs of IS in male patients aged 25–39 years and over 80 years were on the rise, while the EAPC of female patients of all ages was negative, indicating that women as a whole had a downward trend for stroke and cases were better controlled than in men (Supplementary Table 1). For stroke mortality trends in China, from 1980 to 2021, IS mortality in men aged 25–39 years and older than 80 years showed an increasing trend, while mortality in women of all ages showed a continuous decreasing trend (Supplementary Table 2). For the other subtypes of stroke (ICH and SAH), the trend of age-standardized rates was basically the same in males and females, and both showed a downward trend (Supplementary Tables 1 and 2).

The ASIR of IS has increased continuously in the past 30 years, but the ASMR and ASDR peaked in 2004, and the overall trend was still downwards (Figure 1 A). For ICH, the ASIR did not change significantly before 2004, but began to decline significantly after 2004; the ASDR and ASMR increased slightly in 2004 and were on a downward trend at other points in time (Figure 1 B). The ASIR of SAH decreased significantly from 1993 to 2014, and then leveled off. The ASMR and ASDR showed a steady downward trend (Figure 1 C). Trends were found to be relatively similar for men and women.

**Figure 1.**
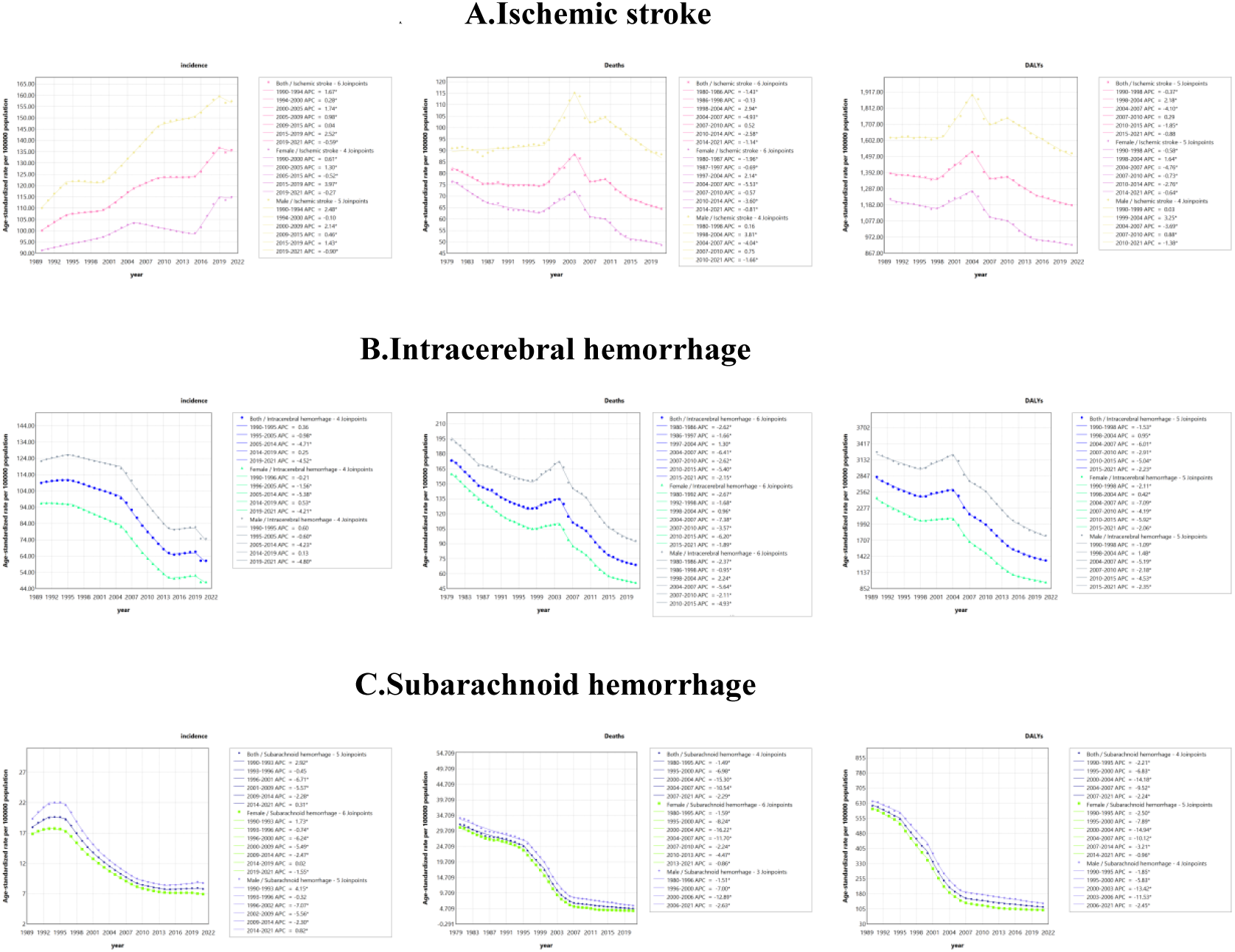
Joinpoint regression analysis of the incidence and DALYs for stroke subtype in China from 1990 to 2021. Death (from 1980 to 2021) (A) IS; (B) ICH; (C) SAH. * Indicates that the Annual Percent Change (APC) is significantly different from zero at the alpha = 0.05 level.

The world is categorized into five groups based on the SDI value. Comparing China with other SDI regions, the incidence, death, and disability associated with IS in China are very high, and the overall control of IS is not as good as that in other SDI regions. The incidence of IS even showed a linear upward trend. The control of ICH and SAH was best in China, exceeding that of other SDI regions (Figure 2).

**Figure 2.**
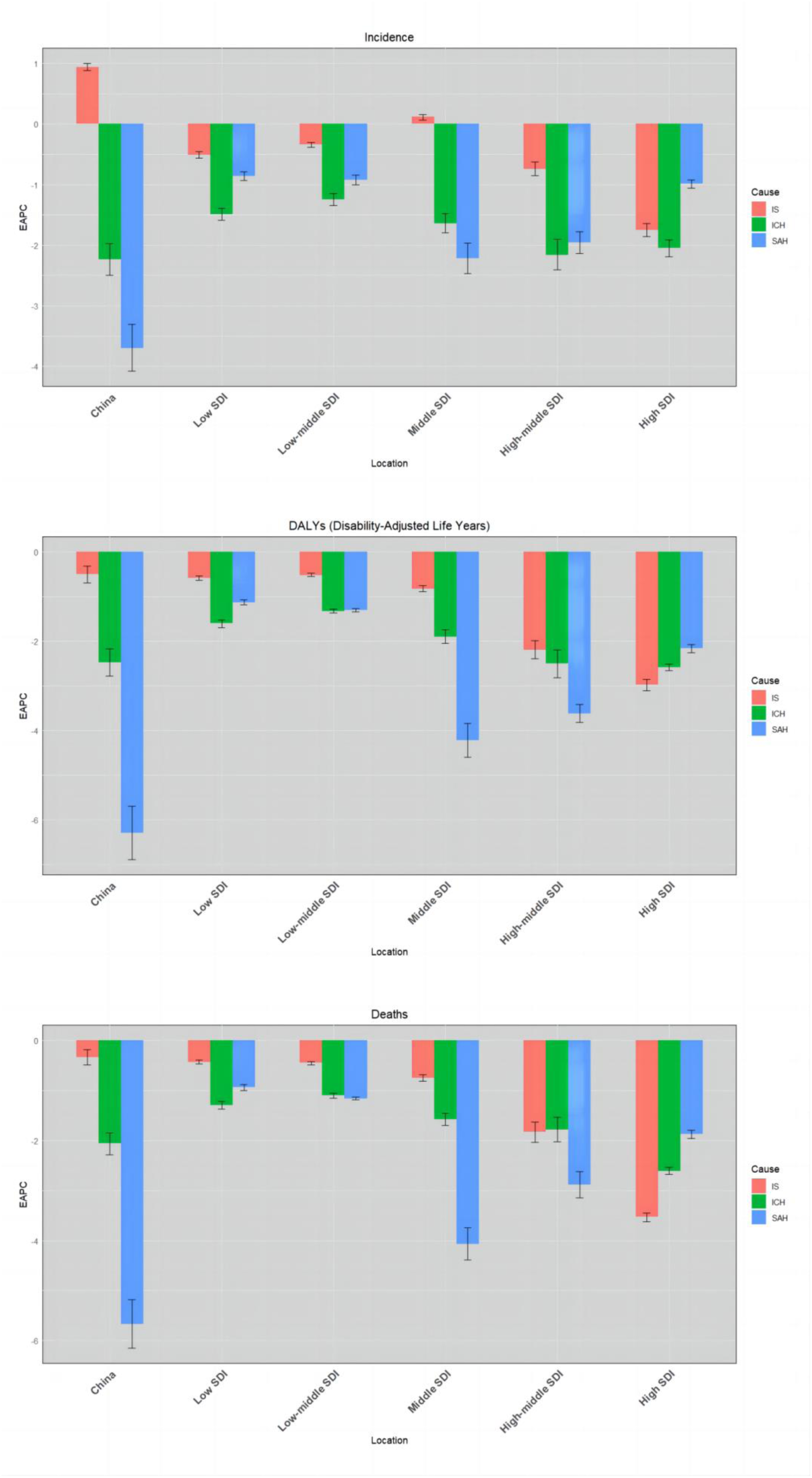
EAPC of ASIR and ASDR for overall stroke trends in China and different SDI regions from 1990 to 2021, disaggregated by sex. ASMR (from 1980 to 2021)

### Population aging in China

From 1990 (100 million) to 2021 (269 million), the number of people aged 60 years and above in China increased by 169%. The incidences, deaths, and DALYs associated with stroke increased by 195% (from 1.07 million to 3.17 million), 90% (from 1.21 million to 2.3 million), and 71% (from 23.69 million to 40.49 million), respectively, in people aged 60 years and older.

Using 1990 as the reference standard, from 1991 to 2021, the incidence and DALYs of IS, ICH, and SAH began to increase each year for both men and women due to population aging and population growth (Figure 3). For mortality rates, 1980 was taken as the reference; from 1981 to 2021, the changes in age-specific mortality rates for IS are correlated with increases in the number of deaths (Figure 3 A). The causes of death from ICH were offset by changes in death rate and the number of deaths due to population aging, resulting in a similar number of deaths from ICH to deaths due to population growth (Figure 3 B). For SAH, the change in age-specific mortality rates is correlated with a decrease in the number of deaths due to SAH, showing a downward trend until 2005, after which it leveled off (Figure 3 C). Trends in morbidity and DALYs related to different stroke subtypes were relatively similar in men and women (Figure 3).

**Figure 3.**
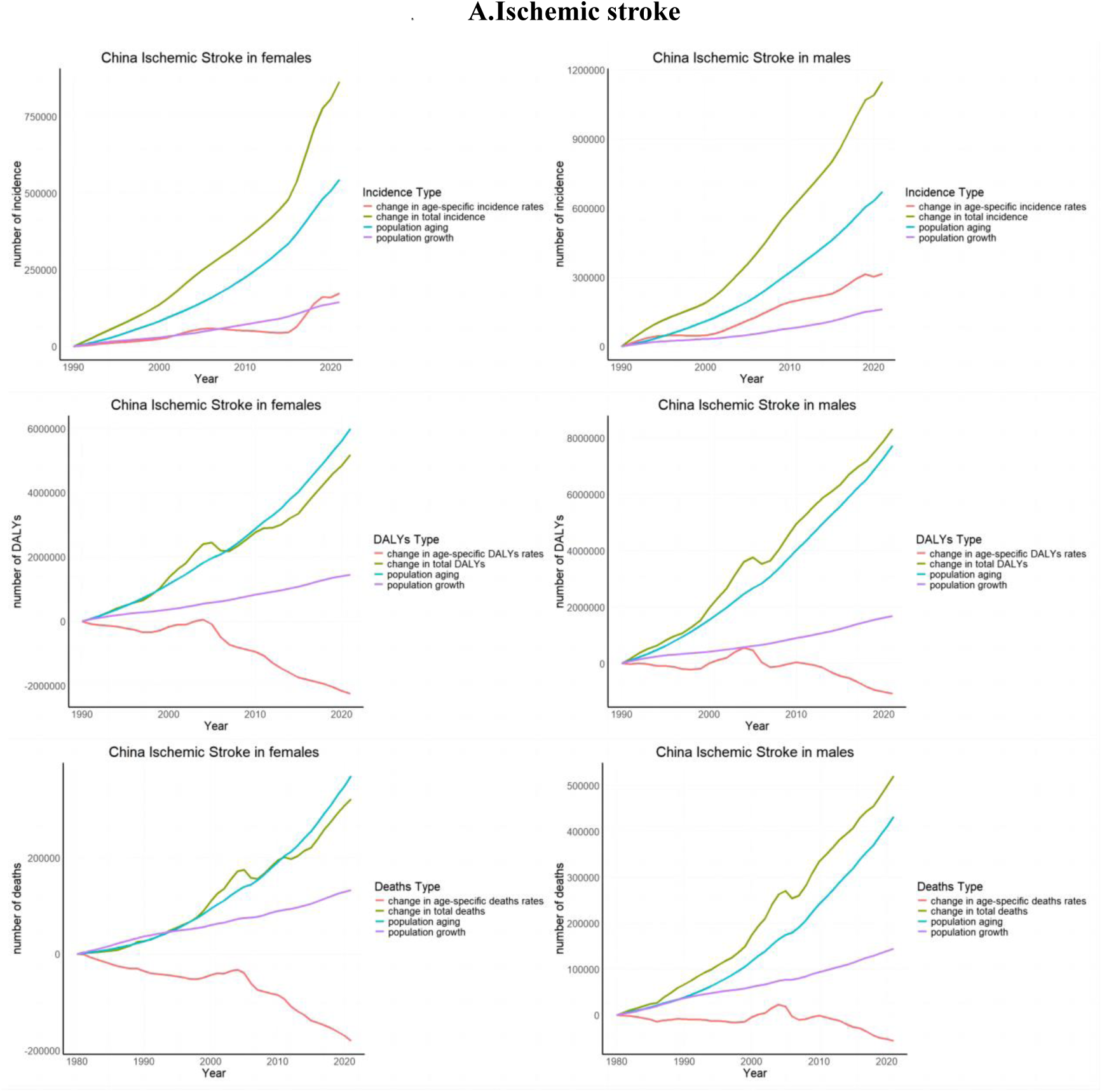
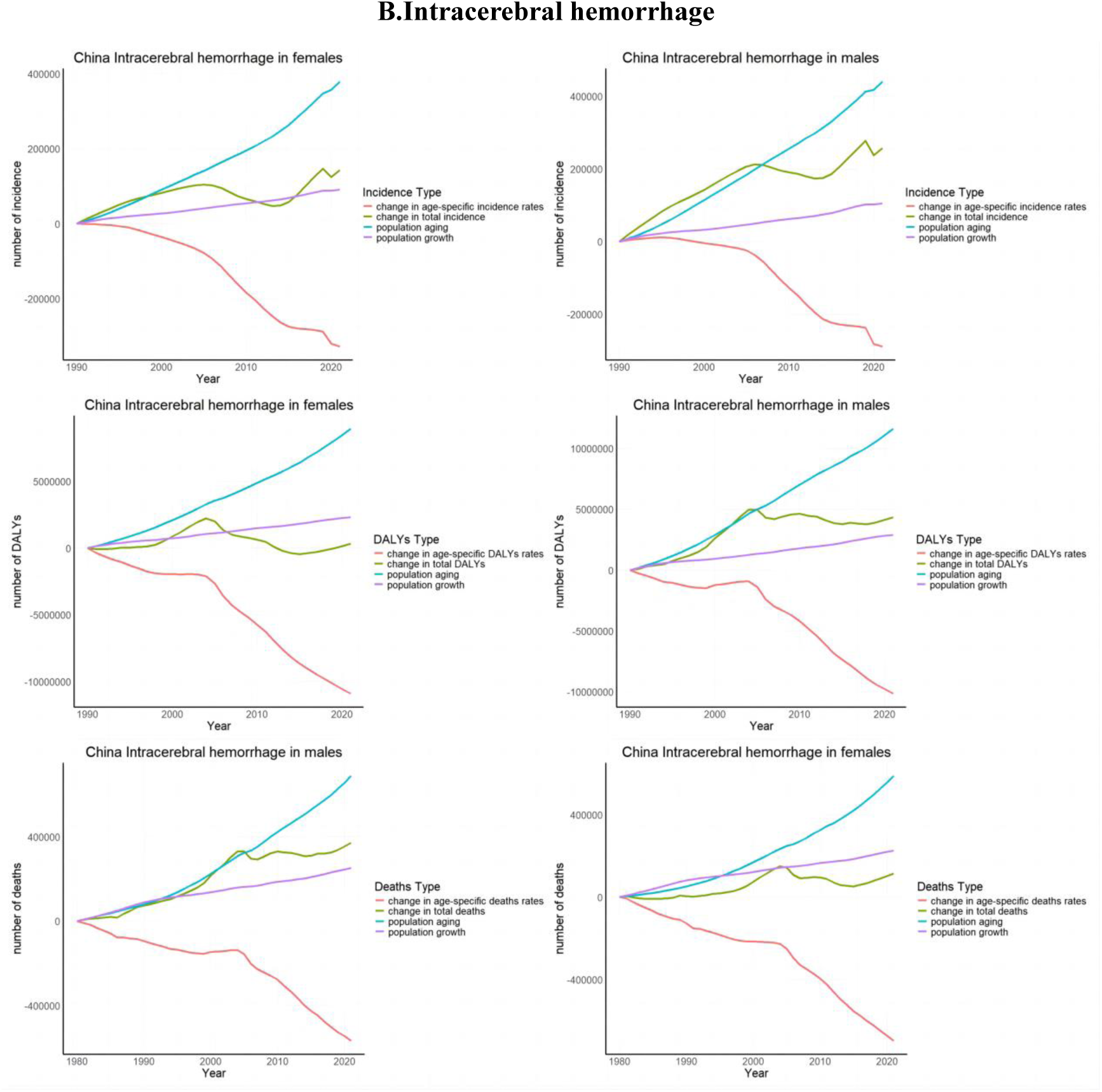
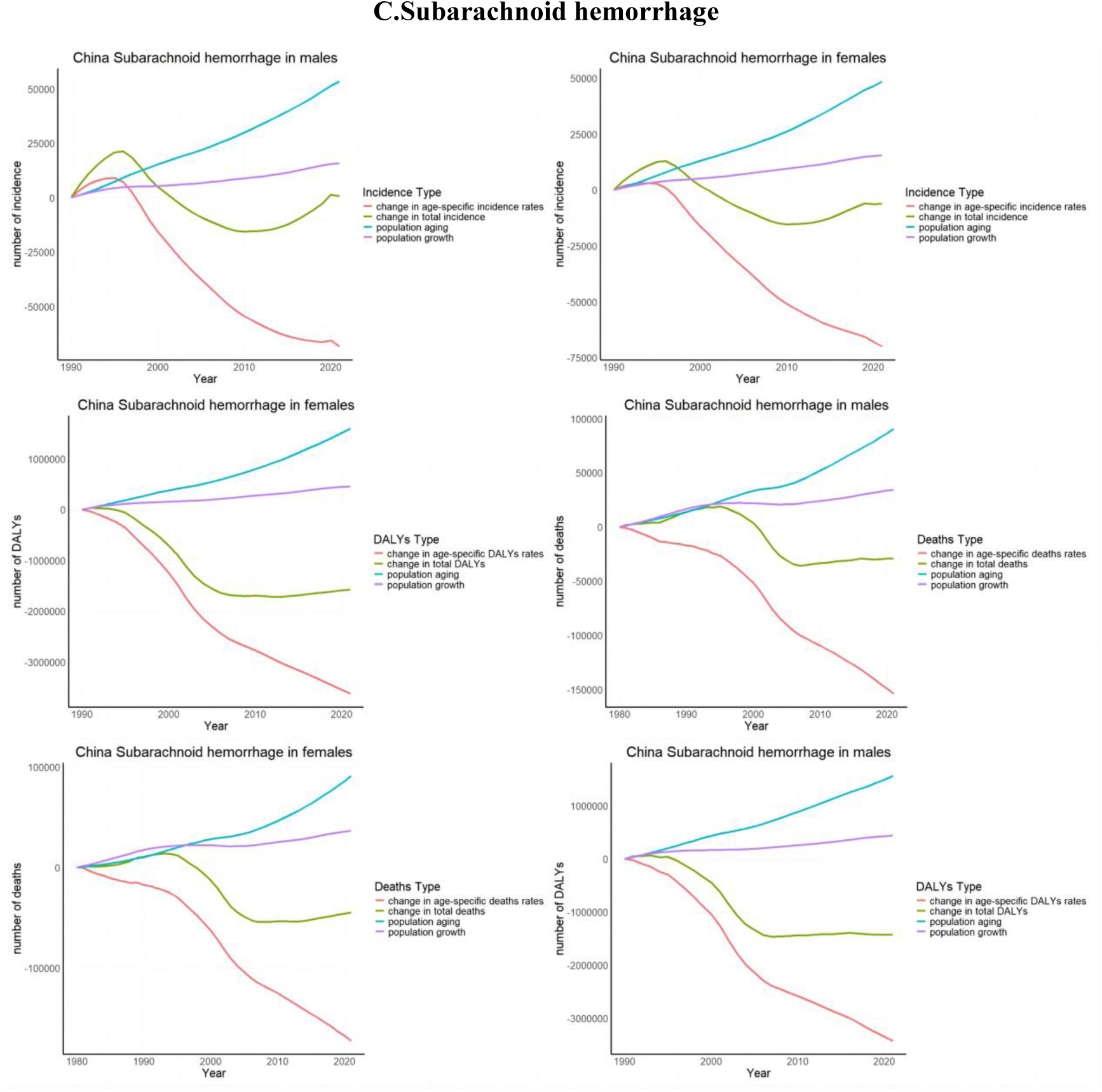
Changes in population aging, population growth, and characteristic age change rates associated with stroke subtype morbidity, disability, and death. (A) IS; (B) ICH; (C) SAH.

Between 1981 and 2021, the proportion of deaths due to population changes in the stroke subtype increased steadily in both men and women (Figure 4). The rate of increase in deaths was higher in the IS group compared to the ICH and SAH groups. The trend was similar among men and women. From 1981 to 2021, the proportion of IS attributed to men was 279.4%, ICH 167.1%, SAH 116% (Figure 4), and the corresponding proportion for women was 204.8%, 138.2%, and 104% (Figure 4).

**Figure 4.**
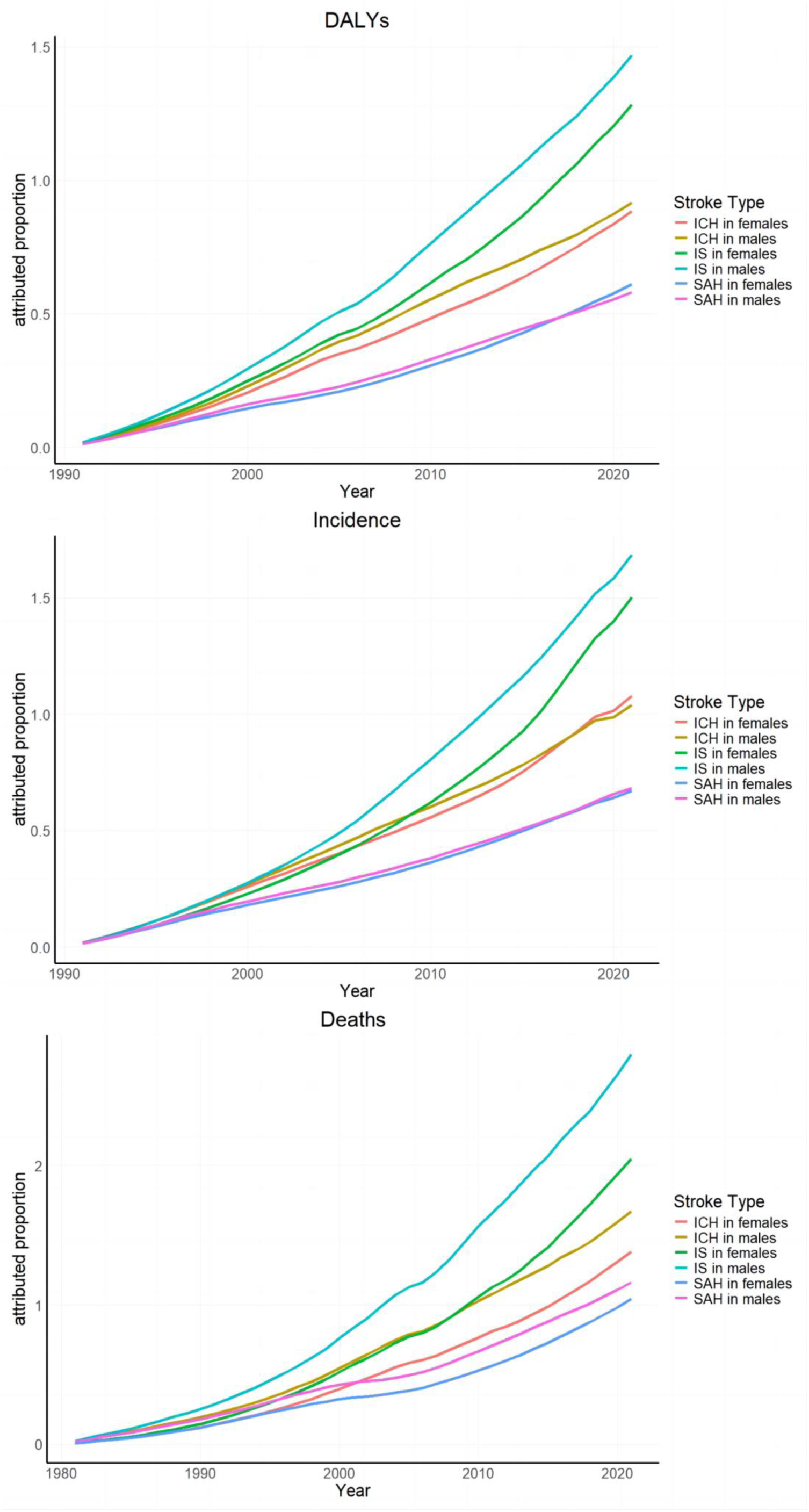
The attributed proportions of stroke incidence, DALYs and death associated with population aging by sex. (A) incidence; (B) DALYs; (C) death.

## Discussion

In our study period, the incidence, deaths, and disability associated with stroke in China and elsewhere in the world were on the rise, while the overall age-standardization trend was on the decline from 1990 to 2021. This may be due to healthcare innovations and socio-economic developments, including stroke screening, emergency green pathways, and early rehabilitation services^[13]^. However, this did not reduce the upward trend, which was largely due to the aging of the population, which has exerted great pressure on society, the economy, and health care. According to the results of China’s seventh census, the population aged 65 years and above in China is 13.5 percent, or about 190 million people^[14]^. An aging population, overall population growth, and a decrease in age-specific rates may be responsible for the increase in stroke prevalence. China is now making use of various policies, institutions, and technical means to promote the management of population aging. However, many young people suffered from early stroke disease, and thus, stroke was not only a disease of the elderly.

The study demonstrated that the ASMR and ASDR for stroke peaked in 2004 and were on a downward trend since then, possibly due to the proclamation of World Stroke Day in 2004 to increase public awareness of stroke risk, prevention, treatment, and rehabilitation through worldwide awareness and education campaigns^[15]^. This initiative could address the challenge of stroke as a major global health problem and promote cooperation and progress in stroke prevention and treatment. Taking China as an example, China’s stroke prevention and treatment work was in line with international standards, learning from and adopting the world’s advanced stroke management and rehabilitation model, which may have more effectively controlled stroke in China after 2004.

The absolute number of stroke cases, deaths, and disabilities is increasing in China, with a much higher growth trend than in Asia and other middle-income countries (South-Asian countries such as India, and developing countries in South-East Asia, such as Indonesia and Malaysia)^[16, 17]^. However, the ASIR decreased in Iran, Japan, and South Korea, which is similar to the situation in China^[18]^. China’s stroke disease burden is similar to those of the middle-SDI countries and high-SDI countries, placing it ahead of developing countries. It can be seen that China’s disease control system was developing in the direction of developed countries, but there are still some challenges and gaps to overcome.

Regarding the differences in stroke subtypes, in our study it was found that IS accounted for an increasing proportion of total strokes, similar to the current research results in the United States, and has become the main focus of the problem^[19]^. The deaths and disability due to IS in the elderly aged over 80 years has been on the rise over the past 40 years. It is worth noting that the incidence of IS is increasing after the age of 20 years, resulting in an increased disease burden for the Chinese people. There has been a significant increase in the absolute number and age-standardized rates of stroke among young people. This increase may be due to cardiovascular risk factors and rising rates of obesity. Several reports indicate a rising incidence of stroke among young people^[20, 21]^. Hence, it is crucial to establish prevention programs aimed at alleviating the burden of stroke in this demographic. ICH and SAH showed downward trends, and SAH showed a significant decline. The Chinese government is actively supporting stroke-related research and clinical practice, promoting the progress of medical technology, and improving the survival rate and quality of life of stroke patients^[22]^. Regarding sex differences, previous studies have demonstrated a higher incidence of stroke in men compared to women^[23–25]^. In our study, although IS was on the rise in both men and women, the magnitude of the trend was lower in women than in men, the decline in ICH and SAH in women was faster than in men, you could see that women had more control than men, which may be caused by differences in social and behavioral backgrounds^[26]^. In 1980, the number of stroke deaths was higher in women than in men, but by 2021 the number of deaths in men had increased significantly; during this period, men still had higher ASMR than women, but the decline in IS, ICH, and SAH was less in men than in women. In this study, IS in men has been shown to be increasing and we should pay attention to this problem. In order to reduce the burden of stroke in China, we should first strengthen awareness of stroke prevention, early identification, and treatment, promote a healthy lifestyle, formulate and implement policies and plans for stroke, improve the level of health care, and implement social support.

The rapid growth of the elderly population underlies stroke deaths in China, which is now moving toward a dual pattern of increasing aging and declining birth rates^[27]^. Stroke remains a challenge in China and could also lead to increased medical costs in the future, significantly increasing the burden on the elderly^[12]^. The burden associated with population aging is largely offset by the reduced burden resulting from prevention and treatment^[28]^. In this study, corresponding data were provided for incidence, death, and DALYs caused by population aging, and it was found that the situation was roughly similar between males and females, but there were great differences among stroke subtypes. Taking ICH as an example, the increase in the number of deaths related to population aging and the decrease caused by its age-specific rate counteracted each other. The trends in incidence and DALYs were relatively similar to those in stroke deaths. It is necessary to introduce better prevention and health management of disease subtypes and improve medical services, so as to enhance the health and quality of life of the elderly.

The main limitations of this study are as follows. First, most of the data in this study were modeled using the DisMod-MR regression tool, and previous research has described the inevitable limitations of the GBD method^[3, 29]^. Second, there was a lack of analysis of provinces in China, and there was no analysis of risk factors associated with stroke. Third, when using the decomposition method, this study only considered the changes of population size, population aging, and age-specific rates, and did not consider the differences in other factors related to the overall rate change. Declining fertility and increasing life expectancy are the two important factors leading to population aging^[6]^, and these mechanisms were not explored.

### Conclusion

This study shows that in recent decades, the incidence, disability, and mortality rates due to ICH and SAH have decreased in both men and women. However, women experience a significantly lower stroke burden than men, while the incidence and mortality rates associated with IS have shown an upward trend. Compared to both developed and developing countries, China has made significant progress in the management and control of ICH and SAH. However, the management of IS remains insufficient and needs further optimization to enhance the overall level of stroke prevention and treatment. The aging population in China contributes to more than half of the increase in stroke burden. To address the growing burden associated with population aging, governments should develop policies to support early screening and intervention measures for stroke, and encourage new technologies and methods for stroke treatment, thereby improving the quality of life of the elderly population.

## Acknowledgements

We appreciate the efforts of the collaborators involved in the 2021 Global Burden of Disease (GBD) study, as well as all colleagues who participated in this research.

## Ethics approval

We utilized data from the publicly accessible GBD 2019 study for this analysis.

## Funding

Not applicable.

## Contributions

Huifeng Liang and Xin Chen conceived and designed the study. Weiwei, Meiying Song, and Huijia Shao extracted the data. Mihriya·Mutallip, XiaoCheng Bao, ShuQun Yang, and Chun Zhang analyzed the data. Huifeng Liang led the manuscript drafting. All authors reviewed the manuscript.

## Competing interests

The authors declare no competing interests.

## Provenance and peer review

Not commissioned; externally peer reviewed.

## Data availability

The data used in this study are derived from the Global Burden of Disease (GBD) database, provided by the Institute for Health Metrics and Evaluation (IHME). The data were accessed through the IHME website (https://www.healthdata.org/gbd), and users are required to register and agree to the data use terms provided by IHME. The GBD database encompasses comprehensive global health metrics, including disease burden, mortality rates, and prevalence. Any publications resulting from this research must adhere to the data use terms and correctly cite the data source. For citation guidelines, please refer to the data use agreement available on the IHME website.

## Notes

### Competing Interest Statement

The authors have declared no competing interest.

### Clinical Trial

The reason for not registering this study is that this was a small-scale exploratory project that did not meet the requirements for mandatory clinical trial registration.

### Author Declarations

We utilized data from the publicly accessible GBD 2019 study for this analysis.

